# Predictors of methadone response in chronic neuropathic pain

**DOI:** 10.64898/2026.10.16.26365778

**Authors:** Marcell Maduro Barbosa, Jorge Alberto Martins Pentiado Júnior, Gabriel Taricani Kubota, Pedro Nascimento Martins, Larissa Iulle Moreira, Ana Mércia Fernandes, Valquíria Aparecida da Silva, Jefferson Rosi Júnior, Eberval Gadelha Figueiredo, Lin Tchia Yeng, Manoel Jacobsen Teixeira, Daniel Ciampi de Andrade

## Abstract

**Background:** Although opioids are frequently prescribed for refractory chronic neuropathic pain, few data are available on baseline factors that may identify individuals more likely to respond and inform individually tailored practice.

**Methods:** This was a post hoc, exploratory analysis of individual-level data from a placebo-controlled trial of methadone for chronic neuropathic pain (NCT05235191). Baseline clinical, psychological, psychophysical and radiological variables were tested for association with ≥30% pain reduction in the methadone group. The correlation between expectation of improvement and pain reduction was examined in both groups.

**Results:** Of the 81 participants completing the 8-week follow-up, 28 (70%) on methadone and 14 (34.1%) on placebo achieved ≥30% pain reduction. In the methadone group, responders had higher baseline expectation of improvement (median 100 [80-100] vs. 55 [50-81]; p=0.004), which correlated moderately with percentage pain reduction (r_s_=0.508; p<0.001). Neither finding was observed in the placebo group (p=0.211 and p=0.161, respectively), although the correlation did not differ significantly between groups (p for interaction=0.728). Other clinical, sociodemographic, psychological and psychophysical variables were not associated with response. In the subsample with peripheral neuropathic pain and structural MRI (n=37), lower left precentral cortex volume was associated with response to methadone (p=0.041), and lower right paracentral volume with response to placebo (p=0.003).

**Conclusions:** Baseline expectation may influence treatment outcomes, supporting its inclusion as a covariate to improve the precision of treatment effect estimates in trials, and its use in clinical practice to help tailor treatment to individual patients. These exploratory findings require confirmation in larger studies.

**Significance statement:** Predictors of opioid response in chronic neuropathic pain remain poorly understood, and treatment expectation has received little attention in this setting. In this study, a single baseline rating of expected improvement identified methadone responders more consistently than pain phenotype, conditioned pain modulation or other psychological traits. These results support considering patient expectation both when interpreting opioid trials and when discussing likely benefit with patients before starting treatment.

## Introduction

Despite ongoing translational and clinical research, treatment for chronic neuropathic pain (NP) remains challenging. Current guideline-supported first-line therapies show only modest efficacy, with number needed to treat (NNT) values ranging from 4.6 to 8.9 (Soliman et al., 2025). Indeed, large prospective real-world studies indicate that fewer than one-third of patients achieve clinically meaningful pain reductions (Moisset et al., 2022; Moulin et al., 2015). In this context, although opioids are recommended only as third-line therapy, reserved for otherwise refractory cases, they are prescribed to 30–50% of patients (Moisset et al., 2022; Moulin et al., 2015).

Particularly, methadone, an atypical opioid with N-methyl-D-aspartate receptor antagonism and serotonin and noradrenaline reuptake inhibition properties, has recently been shown to be effective and well-tolerated for treating NP in an 8-week randomized placebo-controlled clinical trial (Pentiado Júnior et al., 2025). In addition, when compared with other opioids, methadone has been reported to present with a more favorable safety profile, a lower propensity to result in opioid-induced hyperalgesia (Kreek, 2000; Trafton and Ramani, 2009), and lower abuse liability (Jordan et al., 2019), being widely used in the treatment of opioid use disorder (Jordan et al., 2019; Kreek, 2000).

Nonetheless, long-term opioid use is associated with important risks. It is estimated that nearly 80% of long-term users experience at least one adverse event, with serious events occurring in 7.5% (Els et al., 2017), and that 1.5-4.7% develop aberrant drug-related behavior (Bialas et al., 2020). Identifying patients who are more likely to respond to these medications is therefore critical to inform individual risk–benefit judgement (Wilder-Smith, 2007). Beyond this, such information may help to avoid prolonged titration in non-responders (Bouhassira et al., 2014; Emir et al., 2017); to define subsequent steps in patients already receiving treatment (Bouhassira et al., 2014); to enable early detection of reversible factors underlying non-response (McCowat et al., 2019; Schütze et al., 2018); and, ultimately, to advance understanding of the mechanisms of chronic pain, nociception, antinociception and endogenous pain modulation (Baron et al., 2009; Goudman et al., 2022).

Although several variables have been proposed to predict treatment response in NP, including clinical (Bouhassira et al., 2014; Demant et al., 2014), psychophysical (Yarnitsky et al., 2012), psychological (McCowat et al., 2019; Schütze et al., 2018), and radiological features (Lins et al., 2025; Vachon-Presseau et al., 2018); they have not been consistently confirmed in clinical trials. Furthermore, data about response prediction specifically to opioids are particularly scarce. The few studies that have addressed this question were restricted to specific peripheral neuropathic pain conditions (Edwards et al., 2006; Gilron et al., 2013), acute opioid analgesia (Dellemijn and Vanneste, 1997), and examined clinical, psychophysical or only genetic variables (Benavides et al., 2020; Dellemijn and Vanneste, 1997; Edwards et al., 2006; Gilron et al., 2013), whereas radiological and psychological predictors remain largely unexplored.

To help address this gap, we have conducted an exploratory *post hoc* analysis of data from the placebo-controlled trial of methadone for NP described above (Pentiado Júnior et al., 2025), to explore baseline clinical, psychological, psychophysical and radiological variables that may be associated with analgesic response.

## Materials and Methods

### Study design, ethical aspects and participants of the parent study

This *post hoc* analysis used patient-level data from previously published a single-center, randomized, double-blind, placebo-controlled trial conducted at the outpatient Pain Center of the Hospital das Clínicas, University of São Paulo (São Paulo, Brazil), between September 2019 and January 2023 (Pentiado Júnior et al., 2025). The parent study was approved by the institutional ethics committee (CAAE 45397515.2.0000.0068) and registered in ClinicalTrials.gov (NCT05235191) before the enrollment of the first participant.

This study included adults aged 18–85 years-old, who fulfilled the IASP criteria for probable or definite neuropathic pain (Finnerup et al., 2016), confirmed by two independent pain specialists; scored ≥4 on the Douleur Neuropathique en 4 questions (DN4); and presented with pain duration ≥ 6 months, present on most days, and of at least moderate intensity (≥40 mm on a 100-mm visual analogue scale, VAS) despite adequate trials of first-and/or second-line therapies (Finnerup et al., 2015). Exclusion criteria included: i. use of strong opioids within 30 days of enrolment; ii. active substance use disorder, comorbidities posing risk to methadone use (e.g., significant hepatic, renal, or cardiac disease; known prolonged QT interval); iii. pregnancy, or lactation. Participants were randomized (1:1) to receive methadone 5 mg twice daily or matching placebo. Both groups underwent a pre-specified titration protocol over three weeks (up to 30 mg/day or the highest tolerated dose), and maintained for another 5 weeks. Ongoing medication regimens remained stable throughout the study. Primary outcome was defined as a ≥ 30% reduction in VAS score for pain intensity at the end of follow-up.

The trial enrolled 86 participants (Table S1) and median daily methadone dose after titration was 15 (10-20) mg in the active group. Methadone was superior to placebo in reducing mean baseline pain intensity (RR: 2.05; 95% CI: 1.32-3.46; p=0.003), pain severity (p<0.001), superficial spontaneous pain (p=0.025) and paroxysmal pain (p=0.005) dimensions, improved quality (p<0.001) and quantity (p=0.005) of sleep, with no significant adverse events (Pentiado Júnior et al., 2025). Data from baseline and the final visits of all participants who completed the 8-week follow-up were included in the present analysis.

### Variables

#### Pain assessment

Pain was classified by the location of the underlying neurological lesion, as central or peripheral. Furthermore, pain intensity, characteristics and interference were assessed with the following instruments:

- Visual Analogue Scale (VAS): 100-mm continuous line anchored by 0 (“no pain”) and 100 (“worst pain imaginable”), used to assess average pain intensity (Delgado et al., 2018; Downie et al., 1978).
- Neuropathic Pain Symptom Inventory (NPSI): assesses five phenotypical dimensions of NP, including burning, pressing (deep), paroxysmal, evoked, and paresthesia/dysesthesia (de Andrade et al., 2011; Bouhassira et al., 2004). It also allows for the classification of NP into three distinct clinical clusters (i.e., deep pain, pinpointed pain, provoked pain) (Bouhassira et al., 2021).
- Interference Scale from Brief Pain Inventory (BPI-I): investigates pain interference on general activity, mood, walking, work, relationships, sleep, and enjoyment of life (Cleeland and Ryan, 1994; Ferreira et al., 2011).

#### Clinical and Sociodemographic Variables

The following baseline variables were evaluated:

- General clinical and demographic data: age, sex and pain duration, topography of lesion resulting in NP (peripheral vs. central).
- Short-Form 6 Dimensions (SF-6D): a preference-based measure of health-related quality of life (HRQoL) derived from the SF-36, covering six dimensions: physical functioning, role limitations (physical and emotional), social functioning, pain, mental health and vitality. The instrument yields a single utility score anchored at 0 (death) and 1 (full health), with higher scores indicating better HRQoL (Brazier et al., 2002; Campolina et al., 2011).
- Medication Quantification Scale Version III (MQS-III): a tool that quantifies the burden of analgesic medication use. Each drug is scored by multiplying a detriment weight, which reflects the potential for harm of its pharmacological class, according to the dosage level. The individual scores are then summed into a total, with higher totals indicating greater medication burden (Harden et al., 2005).

#### Psychological variables

- Expectation of improvement: Patient expectations with treatment results were measured on a visual analogue scale, anchored at 0 (no improvement) to 100mm (complete pain relief).
- Hospital Anxiety and Depression Scale (HADS) a 14-item self-report questionnaire that assesses anxiety and depressive symptoms. The total score ranges from 0 to 42, with higher scores indicating greater symptom severity. The instrument comprises two 7-item subscales, HADS-A (anxiety) and HADS-D (depression), each scoring from 0 to 21 (Botega et al., 1995; Zigmond and Snaith, 1983).
- Pain Catastrophizing Scale (PCS): a 13-item self-report questionnaire, with each item rated from 0 to 4, yielding a total score from 0 to 52, with higher scores indicating greater catastrophizing. It comprises three subscales: rumination, magnification and helplessness. A total score ≥30 indicates clinically relevant catastrophizing (Sehn et al., 2012; Sullivan and Bishop, 1995).
- Toronto Alexithymia Scale (TAS-20): a 20-item self-report questionnaire, with each item rated from 1 to 5, yielding a total score from 20 to 100, with higher scores indicating greater alexithymia. Scores ≥61 indicate alexithymia, scores from 52 to 60 indicate possible alexithymia, and scores ≤51 indicate absence of alexithymia (Bagby et al., 1994; Yoshida, 2007).

#### Pressure pain threshold (PPT) and Conditioned Pain Modulation (CPM)

Conditioned Pain Modulation (CPM) was assessed using the Cold Pressor Test. Briefly, after establishing the pressure pain threshold (PPT) with an algometer near the tragus (in Newtons/cm²), a 2-seconds suprathreshold stimulus (20% above the threshold) was applied before and immediately after a conditioning stimulus (immersion of the contralateral hand in 4°C water until discomfort reached a level of 6 points on a 0-10 scale). CPM was reported as the difference between VAS scores for pain intensity reported during suprathreshold stimuli before and after the cold pressor test (Yarnitsky et al., 2012). Also, the PPT itself and VAS score reported during baseline suprathreshold stimulus at the tragus were also analyzed as potential predictors.

#### Brain structural measurements

Available baseline structural MRI data from participants with peripheral neuropathic pain and no brain lesions were included in the present analysis. MRI was acquired on a 1.5 T scanner (Magnetom Symphony; Siemens, Erlangen, Germany) equipped with an 8-channel head coil. T1-weighted images were acquired with a 3D Turbo Field Echo (TFE) sequence using the following parameters: repetition time (TR) = 11.51 ms; echo time (TE) = 2.8 ms; matrix size = 288 × 288; slice thickness = 0.9 mm; flip angle = 10°; number of averages = 1.

All T1-weighted images were processed using FreeSurfer software, version 6.0 (Fischl, 2012; Laboratory for Computational Neuroimaging, Athinoula A. Martinos Center for Biomedical Imaging, Charlestown, MA, USA; http://surfer.nmr.mgh.harvard.edu). Automated segmentation was performed using the standard recon-all processing stream (http://surfer.nmr.mgh.harvard.edu/fswiki/recon-all).

Cortical parcellation was performed using the automated Desikan–Killiany–Tourville (DKT) method, which is based on the Desikan–Killiany atlas (Desikan et al., 2006) and was refined by Klein and Tourville (Klein and Tourville, 2012). This method subdivides the cerebral cortex into standard gyral-based neuroanatomical regions suitable for both morphometric and functional analyses.

Cortical regions involved in nociception or antinociception (i.e., the insula, precentral and postcentral gyri, paracentral lobule, caudal anterior cingulate, rostral anterior cingulate, posterior cingulate and isthmus cingulate) were selected as regions of interest. Subsequently, it was investigated whether pain laterality correlated with these measurements (i.e., if pain in the left side of the body correlated with cortical volume in the corresponding region of the contralateral hemisphere, and vice versa).

#### Data analysis strategy

Continuous variables were described as medians (1st and 3rd quartiles) and categorical variables as absolute and relative frequencies. In the methadone group, baseline pain-related, clinical, sociodemographic, psychological, psychophysical and MRI variables were compared between participants with and without a ≥30% pain intensity reduction at the 8-week follow-up visit (primary endpoint) using the Wilcoxon rank sum test (continuous variables) and Fisher’s exact test (categorical variables). In the methadone group, the Wilcoxon rank sum test was also used to compare MRI measurements of the studied brain regions according to pain laterality. Additionally, the relationship between baseline treatment expectation and proportional pain reduction at the final assessment was explored using Spearman’s correlation within each treatment group. Proportional pain reduction was defined as: (baseline VAS score – final VAS score)/ baseline VAS score. The interaction p-value was obtained from a pooled linear regression model of day-56 VAS including treatment, expectation, their interaction, and baseline VAS. Univariate linear regression models were also used to investigate the relationship between VAS pain intensity scores at the final follow-up assessment and (i) cortical volumes of the studied regions in the methadone group and (ii) baseline SF-6D scores in the pooled sample. For the latter, treatment-by-score interaction was assessed in a pooled linear regression model with day-56 VAS as the outcome, adjusting for baseline VAS. Complete cases were analyzed for each variable. A two-sided significance level of 0.05 was adopted for all analyses. All analyses were performed in R, version 4.4.2 (R Core Team, 2024).

## Results

### Participants

Data from the 81 trial participants (methadone n = 40, and placebo n = 41) who completed the 8-week follow-up were included in the analyses. This sample was composed mostly of men (n=49, 60.5%), with median 50 [44,59] years-old. Median baseline pain intensity was 80 [70,90] mm, and median pain duration at enrolment was 6 [3,12] years. Other baseline general demographic, clinical, pain-related and neurophysiological characteristics, according to allocation group, are presented in Table 1. A reduction ≥ 30% in pain intensity was achieved by 28 (70%) of individuals receiving methadone, and 14 (34.1%) of those treated with placebo.

**Table 1.** Baseline characteristics of chronic neuropathic pain patients treated with methadone or placebo, according to their response.

|  | Responder<br>(n = 42) | Non-responder<br>(n = 39) | p |
| --- | --- | --- | --- |
| Age | 54 [48-60] | 48 [43-59] | 0.140 |
| Female<sup>A</sup> | 17 (40%) | 15 (38%) | 0.853 |
| Expectation of improvement | 100 [80-100] | 80 [55-100] | 0.019* |
| Pain duration (years) | 8 [3-15] | 6 [4-12] | 0.547 |
| HADS-A | 6 [3-9] | 7 [4-9] | 0.501 |
| HADS-D | 4 [1-10] | 5 [2-10] | 0.512 |
| HADS Total | 11 [4-17] | 14 [6-19] | 0.418 |
| BPI-I | 73 [53-86] | 63 [50-75] | 0.119 |
| NPSI |  |  |  |
| Burning | 80 [50-90] | 70 [50-80] | 0.291 |
| Pressure | 50 [20-76] | 50 [24-78] | 0.992 |
| Paroxistic pain | 55 [40-89] | 50 [39-83] | 0.528 |
| Evocade pain | 45 [24-66] | 33 [24-67] | 0.663 |
| Parestesia/disesthesia | 80 [51-90] | 65 [50-75] | 0.034* |
| NPSI clusters<sup>A</sup> |  |  | 0.436 |
| Deep pain | 19 (45%) | 23 (59%) |  |
| Pinpointed pain | 16 (38%) | 12 (31%) |  |
| Provoked pain | 7 (17%) | 4 (10%) |  |
| MQS | 18 [13-26] | 23 [17-29] | 0.050 |
| CPM | -10 [-20-0] | -10 [-25-0] | 0.855 |
| Pressure Pain Threshold | 54 [34-72] | 54 [36-69] | 0.817 |
| Suprathreshold VAS Pain Intensity | 50 [30-60] | 50 [20-60] | 0.671 |
| Pain Catastrophism Scale |  |  |  |
| Hopelessness | 11 [6-16] | 10 [6-13] | 0.495 |
| Magnification | 5 [1-8] | 3 [1-6] | 0.187 |
| Rumination | 13 [8-16] | 12 [9-14] | 0.329 |
| Total | 30 [16-37] | 25 [18-32] | 0.202 |
| Alexithymia |  |  |  |

|  | Responder<br>(n = 42) | Non-responder<br>(n = 39) | p |
| --- | --- | --- | --- |
| Factor 1 | 35 [22-45] | 32 [24-42] | 0.548 |
| Factor 2 | 12 [10-15] | 14 [11-16] | 0.202 |
| Factor 3 | 10 [8-13] | 12 [10-15] | 0.083 |
| Total | 67 [58-80] | 70 [56-83] | 0.843 |

*Treatment response was defined as a $\geq 30\%$ reduction in average pain intensity at the 2-month follow-up.*
*Variables presented as median [Q1, Q3], unless otherwise specified.*
<sup>A</sup>Data presented as n (%)
\* $p < 0.05$

### Clinical and Sociodemographic Variables

In the methadone group, no baseline clinical or sociodemographic variables were associated with a ≥ 30% pain reduction at the end of follow-up (Table 2). The presence of allodynia and the location of the causative lesion also did not differ between responders and non-responders (p=0.245 and p=0.612, respectively, Table S2), nor was HRQoL correlated with this outcome (p=0.798, Table S3). Meanwhile, in the pooled sample, including individuals from both the methadone and placebo groups, responders presented at baseline with significantly higher paresthesia/dysesthesia (80 [51-90] vs. 65 [50-75], p=0.034), and a trend for lower MQS scores (18 [13-26] vs. 23 [17-29], p=0.050).

**Table 2.** Baseline characteristics of chronic neuropathic patients treated with methadone according to their response category.

|  | Responder<br>(n = 28) | Non-responder<br>(n = 12) | p |
| --- | --- | --- | --- |
| Age | 55 [48-60] | 50 [43-61] | 0.545 |
| Female | 8 (29%) | 4 (33%) | >0.999 |
| Expectation of improvement | 100 [80-100] | 55 [50-81] | 0.004* |
| Pain duration (Years) | 7 [3-12] | 8 [5-12] | 0.468 |
| HADS-A | 5 [3-8] | 7 [5-8] | 0.374 |
| HADS-D | 4 [1-6] | 8 [3-12] | 0.177 |
| HADS Total | 10 [3-14] | 15 [7-20] | 0.156 |
| BPI-I | 69 [48-82] | 67 [63-79] | 0.802 |
| NPSI |  |  |  |
| Burning | 70 [50-85] | 55 [38-80] | 0.236 |
| Pressure | 50 [0-73] | 33 [15-58] | 0.484 |
| Paroxistic pain | 50 [34-86] | 59 [39-90] | 0.667 |
| Evocade pain | 33 [22-64] | 32 [26-67] | 0.905 |
| Parestesia/dysesthesia | 70 [50-91] | 61 [49-76] | 0.285 |
| NPSI clusters |  |  | >0.999 |
| Deep pain | 14 (50%) | 6 (50%) |  |
| Pinpointed pain | 10 (36%) | 4 (33%) |  |
| Provoked pain | 4 (14%) | 2 (17%) |  |
| MQS | 20 [13-26] | 20 [17-26] | 0.585 |
| CPM | -15 [-20-0] | -10 [-30-0] | 0.928 |
| Pressure Pain Threshold | 63 [46-79] | 62 [41-298] | 0.525 |
| Suprathreshold VAS Pain Intensity | 50 [30-63] | 50 [40-60] | 0.964 |
| Pain Catastrophism Scale |  |  |  |
| Hopelessness | 10 [6-15] | 11 [10-15] | 0.801 |
| Magnification | 4 [1-7] | 3 [2-3] | 0.542 |
| Rumination | 13 [8-15] | 12 [9-13] | 0.381 |
| Total | 27 [15-34] | 26 [22-32] | 0.723 |
| Alexithymia |  |  |  |

|  | Responder<br>(n = 28) | Non-responder<br>(n = 12) | p |
| --- | --- | --- | --- |
| Factor 1 | 31 [21-42] | 29 [21-32] | 0.352 |
| Factor 2 | 12 [11-17] | 15 [11-17] | 0.604 |
| Factor 3 | 10 [8-12] | 13 [11-15] | 0.196 |
| Total | 67 [58-77] | 61 [57-73] | 0.442 |

*Treatment response was defined as a $\geq 30\%$ reduction in average pain intensity at the 2-month follow-up.*
*Variables presented as median [25%-75%], unless otherwise specified.*
<sup>A</sup>Data presented as n (%)
\* $p < 0.05$

### Psychological Variables

Participants achieving ≥ 30% pain reduction presented with significantly higher expectation of improvement with treatment at baseline, both in the pooled sample (100 [80-100] vs. 80 [55-100], p=0.019, Table 1), and the methadone group (100 [80-100] vs. 55 [50-81], p=0.004, Table 2). However, in the placebo group, expectation levels were not associated with pain improvement (p=0.211, Table S4). Additionally, baseline expectation levels also correlated moderately with proportional pain reduction at the last follow-up (r_s_ = 0.508, p < 0.001, Table 3) among individuals receiving methadone, but not in the placebo group (p = 0.161). Nonetheless, interaction analysis did not find evidence of significant difference of correlation between baseline expectation levels and proportional pain reduction between treatment groups (p=0.728, Table 3). Anxiety, depression, alexithymia and pain catastrophism scores did not differ significantly between responders and non-responders in the pooled sample and methadone group (Tables 1 and 2).

**Table 3.** Correlation between expectation of improvement and treatment response according to group.

| Group | $r_s$ | p-value | p interaction |
| --- | --- | --- | --- |
| Methadone | 0.508 | <0.001* | 0.728 |
| Placebo | 0.223 | 0.161 |  |

*Spearman Rank Correlation*
\* $p < 0.05$

### Psychophysical Variables

Among participants receiving methadone, baseline CPM (p = 0.855), PPT (p = 0.817), and suprathreshold VAS intensity (p = 0.817) were not significantly associated with ≥30% pain reduction at the end of follow-up. Similarly, none of these variables was associated with this outcome in the pooled sample.

### Structural cortical variables

Of the participants who completed the 8-week follow-up, 37 (methadone n = 15; placebo n = 22) had peripheral neuropathic pain, no brain lesions, and available baseline structural brain MRI (Table S5). In the methadone group, 13 participants (86.7%) achieved ≥30% pain reduction. Compared with non-responders, responders had a lower median left precentral cortex volume (10,507 [9,963-11,620] vs. 13,414 [12,966-13,861] voxels; p = 0.041). No other cortical region was associated with response (Table S6). Furthermore, no cortical region was significantly correlated with pain intensity at the 8-week follow-up, except for a borderline association between larger right posterior cingulum volume and higher pain intensity (β = 3.9, 95% CI 0.0-7.8; Table S7).

In the placebo group, 8 participants (36.4%) were responders. Responders had a lower median right paracentral cortex volume than non-responders (2,791 [2,709-2,945] vs. 3,450 [3,034-3,688] voxels; p = 0.003). No other cortical region was associated with response (Table S8).

## Discussion

In this exploratory post hoc analysis of METHA-NeP, a randomized, placebo-controlled trial of methadone for chronic neuropathic pain, higher baseline expectation of improvement was the most consistent predictor of response. Among participants receiving methadone, responders reported higher baseline expectation than non-responders, and expectation correlated moderately with the magnitude of pain relief. Neither association was significant in the placebo group. However, the correlation between expectation and pain reduction did not differ significantly between groups (p for interaction = 0.728). Therefore, it is not possible to establish that the association is specific to the active drug, and an influence of expectation regardless of treatment cannot be excluded. By contrast, baseline clinical, sociodemographic, other psychological and psychophysical measures, including CPM and PPT, did not distinguish responders from non-responders. In the MRI subsample, lower left precentral and lower right paracentral cortex volumes were associated with response in the methadone and placebo groups, respectively.

Evidence on predictors of opioid analgesia in neuropathic pain is limited to a few small secondary analyses of clinical trials. Among demographic and clinical variables, younger age and higher baseline pain intensity predicted opioid response in one study (Edwards et al., 2006), whereas age, sex and pain duration were not associated with response in another (Gilron et al., 2013), as in our sample. Regarding pain phenotype, the presumed mechanism of neuropathic pain did not predict response to a single 5-hour fentanyl infusion (Dellemijn and Vanneste, 1997), consistent with the absence of association with lesion topography (central vs. peripheral) in our study, although these classifications are not equivalent. Moderate-to-severe “throbbing” pain predicted a poorer response to morphine, whereas allodynia did not (Gilron et al., 2013). Likewise, we found no association for allodynia, nor for any NPSI dimension or cluster. Psychophysically, higher heat pain thresholds predicted greater opioid analgesia (Edwards et al., 2006), whereas pressure pain thresholds did not in our study. However, these modalities engage distinct afferent pathways and may not behave similarly. Finally, no candidate polymorphism predicted response to morphine monotherapy (Benavides et al., 2020). None of these studies assessed other psychological factors or neuroimaging, nor measured treatment expectation.

Nonetheless, expectation of improvement has increasingly been recognized as a determinant of outcome in chronic pain treatment. Expectations are considered a central component of placebo analgesia (Lunde et al., 2024), and baseline expectations have also been associated with outcomes of active interventions (Langford et al., 2023). For instance, higher expectations predicted improvement after chiropractic care for low back pain (Eklund et al., 2019), and after multidisciplinary treatment in a real-world cohort of 2,272 patients with chronic pain (Cormier et al., 2016). Most of these studies, however, investigated acupuncture or physical therapies for musculoskeletal disorders, and assessed expectations heterogeneously, mostly through unvalidated, variably worded single items (Langford et al., 2023). Notably, although expectations may influence the response to both placebo and active treatments, their effect may not be similar. In a crossover trial in chronic low back pain, expectations predicted analgesia after verum, but not sham acupuncture (Wasan et al., 2010). Similarly, in a trial of amitriptyline versus active placebo for chronic pain after spinal cord injury, expectation of relief correlated with pain reduction only with amitriptyline (r=0.37 vs. −0.16), with a significant treatment-by-expectation interaction (Turner et al., 2002). Conversely, experimental manipulations of expectation have produced larger effects in placebo than in active treatment groups (Langford et al., 2023). In our study, the correlation between expectation and pain reduction was stronger in the methadone (r_s_=0.508) than in the placebo group (r_s_=0.223). Although the interaction was not significant, and the study was not powered to detect it, this pattern is consistent with the findings above and may suggest a greater contribution of expectation to pain relief in the active arm.

Several mechanisms may explain why expectation would be more closely associated with outcome in the active arm. Participants with higher expectations may be more willing to continue treatment despite mild adverse effects, adhere more closely to dosing and titration, use non-pharmacological co-interventions more often, and complete follow-up, all of which are likely to matter more when a pharmacologically active drug is administered. Conversely, negative expectations may amplify the burden of adverse effects. In a randomized trial of a topical analgesic for chronic musculoskeletal pain, high expectations of side effects were associated with more adverse events and lower response rates, particularly in the active arm (Yang et al., 2024). In addition, adverse effects of the active drug may partially unblind participants (Turner et al., 2002), confirming prior expectations and amplifying their influence on reported outcomes (Langford et al., 2023). Regardless of the mechanism, these findings support conducting prespecified adjustment for baseline expectation in analyses of chronic pain trials, which may increase the precision of treatment effect estimates and, consequently, assay sensitivity (Langford et al., 2023). Expectation may also help to choose between treatments with similar supporting evidence. In a randomized trial of cognitive behavioral therapy, exercise, or both for chronic widespread pain, participants who expected improvement from their allocated treatment were more likely to report a positive outcome at the end of treatment (adjusted OR=2.03) and 3 months later (adjusted OR=2.31), whereas being allocated to their preferred treatment had no effect (Beasley et al., 2017).

Meanwhile, the neuroimaging findings should be interpreted with caution. Only two methadone-treated participants were non-responders, multiple regions were tested without correction for multiple comparisons, and no formal treatment-by-region interaction was assessed; therefore, they should be considered hypothesis-generating. Evidence on neuroimaging predictors of response to active analgesics is scarce. In fibromyalgia, baseline insular glutamate and connectivity predicted response to pregabalin, but not placebo (Harris et al., 2013), and resting-state connectivity of the posterior cingulate and dorsolateral prefrontal cortices discriminated pregabalin from milnacipran responders (Ichesco et al., 2021). In knee osteoarthritis, right parahippocampal connectivity was associated with placebo-corrected response to duloxetine (Tétreault et al., 2016). For opioids, the only available evidence comes from healthy volunteers, in whom baseline reward-circuitry responses to experimental pain and trait reward responsiveness predicted remifentanil analgesia (Wanigasekera et al., 2012). All these studies were small, relied on functional rather than structural measures, and none addressed neuropathic pain or sustained opioid treatment.

Interestingly, both regions associated with response in our study belong to the sensorimotor cortex. This resembles the thicker sensorimotor cortex observed in placebo non-responders with chronic back pain (Vachon-Presseau et al., 2018), and suggests that lower sensorimotor grey matter may be associated with a propensity to improve that is not specific to the active drug, in line with our expectation findings. However, cortical volume and thickness are related but distinct measures, and the images from the present study were acquired at 1.5 T. Moreover, the most consistent imaging predictors of placebo response have involved prefrontal regions, such as medial prefrontal–insular connectivity (Hashmi et al., 2012) and right middle frontal gyrus connectivity (Tétreault et al., 2016). Indeed, prefrontal regions, including the dorsolateral, ventromedial and orbitofrontal cortices, are consistently engaged by expectations of pain relief (Atlas and Wager, 2014), but were not among our regions of interest. Future trials combining expectation measures with structural and functional imaging, including prefrontal regions, may help to separate drug-specific from non-specific predictors of response.

This study has several limitations. First, this was an exploratory *post hoc* analysis of a trial not designed or powered to identify predictors of response, and multiple variables were tested without correction for multiple comparisons; therefore, false-positive findings cannot be excluded. Second, the high response rate in the methadone group resulted in only 12 non-responders, limiting power to detect associations. Third, expectation scores showed a ceiling effect, particularly in the placebo group, and the resulting restricted range may have attenuated the correlation observed in this group. Fourth, only participants who completed follow-up were analyzed and, if expectation influenced adherence or retention, this may have introduced selection bias. Fifth, psychophysical assessment was limited to pressure stimuli at a single site, and thermal thresholds were not assessed. Finally, methadone plasma levels and pharmacogenetic factors relevant to its metabolism and action were not assessed, despite the marked interindividual variability in methadone pharmacokinetics and the range of doses achieved.

In conclusion, higher baseline expectation of improvement was the most consistent predictor of analgesic response to methadone in refractory chronic neuropathic pain, whereas other clinical, psychological and psychophysical variables were not. Although the association between expectation and pain relief appeared stronger in the methadone group, it could not be shown to be specific to the active drug. These findings support the routine measurement of baseline expectation, using validated instruments, in chronic pain trials, and its prespecified inclusion as a covariate to improve the precision of treatment effect estimates. In clinical practice, assessing expectations may also help to tailor treatment to the individual patient, particularly when choosing among alternatives with similar evidence of efficacy. Larger prospective studies are needed to confirm these findings and to determine whether expectation moderates the response to opioids.

## Data Availability

All data produced in the present study are available upon reasonable request to the authors.

## Acknowledgements

We thank all the patients who generously participated in this study. We are also grateful to the clinical and research staff of the Pain Center, Department of Neurology, and the Department of Physiotherapy at the University of São Paulo for their invaluable assistance in patient recruitment, data collection, and technical support. The authors used Claude (Anthropic) to assist in drafting and editing of this manuscript. The tool was not used to collect, analyze or interpret data. All AI-assisted text was reviewed, revised and approved by the authors, who take full responsibility for the content of the publication.

## Data Availability

Anonymized data supporting the main findings can be made available to qualified investigators upon reasonable request to the corresponding author.

## Author contributions

This study was designed by DCA and data collection was performed by MMB, JAMP Jr, LIM, AMF and VAS. Data was analyzed and interpreted by MMB, GTK, PNM, DCA, JR Jr, EGF, LTY, and MJT. The draft of the original manuscript was produced by MMB, GTK, and DCA. All authors have discussed the results, commented on the manuscript, and approved it in its final form.

