## Supplementary Tables for "Predictors of methadone response in chronic neuropathic pain"

**Supplementary materials**

Summary

1. **Supplementary Tables**

| **Table S1.** Descriptive analysis from baseline data of the sample | | |
| --- | --- | --- |
| Characteristic | Placebo, N = 42 | Methadone, N = 44 |
| Sex^A^ |  |  |
| Male | 22 (52%) | 29 (66%) |
| Female | 20 (48%) | 15 (34%) |
| Age | 49 (43 – 58) | 54 (44 – 59) |
| Expectation of improvement | 100 (80 – 100) | 83 (50 – 100) |
| Visual Analogue Scale (VAS) | 80 (70 – 90) | 80 (70 – 90) |
| MQS III | 21 (14 – 26) | 18 (13 – 26) |
| HADS-A | 7 (3 – 10) | 6 (3 – 8) |
| HADS-D | 6 (3 – 10) | 4 (1 – 10) |
| HADS - total | 14 (6 – 19) | 10 (4 – 17) |
| Alexithymia - Factor 1 | 37 (26 – 47) | 29 (20 – 40) |
| Alexithymia - Factor 2 | 13 (9 – 15) | 14 (11 – 17) |
| Alexithymia - Factor 3 | 12 (8 – 13) | 12 (8 – 14) |
| Alexitimia categories^A^ |  |  |
| Alexithymia | 29 (73%) | 28 (65%) |
| Possible alexithymia | 7 (18%) | 9 (21%) |
| Non-alexithymia | 4 (10%) | 6 (14%) |
| Catastrophism - Helplessness | 10 (6 – 15) | 10 (6 – 15) |
| Catastrophism - Magnification | 5 (2 – 8) | 3 (1 – 6) |
| Catastrophism - Rumination | 13 (9 – 16) | 12 (5 – 14) |
| Total Catastrophism | 27 (18 – 35) | 26 (15 – 33) |
| NPSI |  |  |
| Burning | 80 (53 – 90) | 70 (50 – 80) |
| Pressure | 58 (29 – 80) | 48 (11 – 66) |
| Paroxistic | 50 (40 – 85) | 50 (35 – 90) |
| Evocated | 47 (27 – 67) | 33 (23 – 67) |
| Paresthesia/dysesthesia | 75 (55 – 85) | 64 (50 – 90) |
| NPSI clusters |  |  |
| Deep pain | 48 (8, 68) | 58 (25, 80) |
| Provoked pain | 33 (23, 67) | 47 (27, 67) |
| Pinpointed pain | 64 (50, 90) | 75 (55, 85) |
| Nervous system lesion topography^A^ |  |  |
| Central | 14 (32%) | 10 (24%) |
| Peripheral | 30 (68%) | 32 (76%) |
| CPM | -10 (-25, 0) | -10 (-30, 0) |
| Pressure Pain Threshold | 59 (41, 80) | 49 (30, 60) |
| Suprathreshold VAS Pain Intensity | 50 (30, 60) | 40 (20, 50) |
| SF-6D | 58 (42, 69) | 54 (23, 62) |
| Allodynia^A^ | 23 (52%) | 18 (43%) |
| Neuropathic pain duration (years) | 8 (4, 13) | 6 (3, 12) |
| *Variables presented as median [25%-75%], unless otherwise specified.*  *^A^Data presented as n (%)* | | |

| **Table S2.** Analysis of allodynia and topography between groups and subgroups | | | | | | |
| --- | --- | --- | --- | --- | --- | --- |
| Variable | Contrast | Group | Difference | IC95% | p-value | P interaction |
| Allodynia | 1 - 0 | Methadone | 8.7738679 | -6.025 a 23.573 | 0.245 | 0.367 |
| Allodynia | 1 - 0 | Placebo | -0.8669538 | -15.602 a 13.868 | 0.908 |  |
| Topography | Periferic - Central | Methadone | -4.0036255 | -19.488 a 11.481 | 0.612 | 0.262 |
| Topography | Periferic - Central | Placebo | -17.1694383 | -33.847 a -0.492 | 0.044 |  |

*VAS differences according to potential predictor variables within treatment allocation groups. Contrasts were obtained using linear regression models with treatment, subgroup, baseline VAS and treatment per subgroup interaction.*

**p < 0.05*

| **Table S3.** Analysis of quality-of-life SF-6D scores and treatment response | | | |
| --- | --- | --- | --- |
| Group | Rho | p-value | P interaction |
| Methadone | -0.042 | 0.798 | 0.555 |
| Placebo | 0.015 | 0.927 |  |

*Pearson correlation coefficient between quality of life and final VAS in placebo and methadone groups.* *Treatment response was defined as a ≥ 30% reduction in average pain intensity at the 2-month follow-up.*

**p < 0.05*

| **Table S4.** Expectation of improvement according to response to treatment in the placebo group | | | |
| --- | --- | --- | --- |
| Characteristic | Responder, N = 14 | Non-responder, N = 27 | p-value |
| Expectation of Improvement |  |  | 0.211 |
| Median | 100 (83, 100) | 90 (80, 100) |  |
| Limits | 60 - 100 | 50 - 100 |  |
| *Wilcoxon Rank Sum Test. Treatment response was defined as a ≥ 30% reduction in average pain intensity at the 2-month follow-up.*  **p < 0.05* | | | |

| **Table S5.** Cortical volume of different brain regions according to treatment group | | |
| --- | --- | --- |
| Region | Methadone  N = 15 | Placebo  N = 22 |
| Insula (L) | 6,292 (5,857, 6,617) | 6,556 (5,815, 6,728) |
| Insula (R) | 6,115 (5,731, 6,551) | 6,250 (5,608, 6,687) |
| Precentral (L) | 10,839 (9,963, 12,170) | 10,819 (9,851, 12,978) |
| Precentral (R) | 11,273 (9,143, 11,905) | 10,367 (9,214, 11,276) |
| Postcentral (L) | 8,012 (7,055, 8,423) | 7,953 (7,308, 8,597) |
| Postcentral (R) | 7,672 (6,727, 8,407) | 7,985 (6,957, 8,462) |
| Paracentral (L) | 3,086 (2,528, 3,310) | 2,858 (2,528, 3,343) |
| Paracentral (R) | 3,387 (2,809, 3,920) | 3,039 (2,849, 3,534) |
| Caudal anterior cingulate (L) | 1,554 (1,294, 1,863) | 1,428 (1,144, 1,607) |
| Caudal anterior cingulate (R) | 1,723 (1,556, 1,889) | 1,570 (1,326, 2,056) |
| Cingular isthmus (L) | 2,461 (2,051, 2,653) | 2,376 (2,123, 2,672) |
| Cingular isthmus (R) | 2,309 (1,916, 2,421) | 2,141 (1,931, 2,381) |
| Posterior cingulate (L) | 2,851 (2,394, 3,071) | 2,591 (2,365, 2,984) |
| Posterior cingulate (R) | 2,771 (2,684, 2,807) | 2,615 (2,364, 3,074) |
| Rostral anterior cingulate (L) | 2,463 (1,819, 2,737) | 2,204 (1,865, 2,574) |
| Rostral anterior cingulate (R) | 1,523 (1,295, 1,828) | 1,633 (1,275, 1,937) |
| *Values presented as median (Q1, Q3).* | | |

| **Table S6.** Cortical volume from different brain regions in the methadone group according to treatment response | | | | | | |
| --- | --- | --- | --- | --- | --- | --- |
|  | Left | | | Right | | |
| Region | Non-responder, N = 2 | Responder, N = 13 | p-value | Non-responder, N = 2 | Responder, N = 13 | p-value |
| Insula |  |  | 0.061 |  |  | 0.089 |
| Median | 6977 (6822, 7131) | 6198 (5857, 6538) |  | 6764 (6657, 6870) | 6111 (5731, 6180) |  |
| Limits | 6667 – 7286 | 5565 – 7286 |  | 6551 – 6976 | 5031 – 6976 |  |
| Precentral |  |  | 0.041* |  |  | 0.061 |
| Median | 13414 (12966, 13861) | 10507 (9963, 11620) |  | 12317 (12158, 12475) | 11244 (9143, 11557) |  |
| Limits | 12519 – 14308 | 8081 – 12519 |  | 12000 – 12633 | 6753 – 12633 |  |
| Post-central |  |  | 0.308 |  |  | 0.865 |
| Median | 9396 (8704, 10087) | 8012 (7055, 8374) |  | 7762 (7717, 7807) | 7672 (6727, 8407) |  |
| Limits | 8012 – 10779 | 6338 – 10113 |  | 7672 – 7852 | 5000 – 9868 |  |
| Paracentral |  |  | 0.174 |  |  | 0.497 |
| Median | 3355 (3333, 3378) | 2961 (2528, 3166) |  | 3669 (3543, 3794) | 3279 (2809, 3920) |  |
| Limits | 3310 – 3400 | 2188 – 3832 |  | 3417 – 3920 | 2408 – 4224 |  |
| Caudal anterior cingulum |  |  | 0.865 |  |  | 0.174 |
| Median | 1510 (1487, 1532) | 1554 (1294, 1863) |  | 2108 (1976, 2240) | 1718 (1556, 1878) |  |
| Limits | 1465 – 1554 | 521 – 2766 |  | 1844 – 2372 | 1326 – 1998 |  |
| Isthmus |  |  | 0.061 |  |  | 0.126 |
| Median | 2751 (2702, 2799) | 2460 (2051, 2476) |  | 2527 (2446, 2609) | 1999 (1916, 2364) |  |
| Limits | 2653 – 2848 | 1748 – 2829 |  | 2364 – 2690 | 1848 – 2545 |  |
| Posterior cingulum |  |  | 0.089 |  |  | 0.089 |
| Median | 3091 (3081, 3100) | 2718 (2394, 2951) |  | 3229 (3018, 3440) | 2768 (2684, 2791) |  |
| Limits | 3071 – 3110 | 1836 – 3157 |  | 2807 – 3651 | 2160 – 3040 |  |
| Rostral anterior cingulum |  |  | 0.308 |  |  | 0.496 |
| Median | 2705 (2584, 2826) | 2193 (1819, 2602) |  | 1747 (1551, 1942) | 1523 (1295, 1692) |  |
| Limits | 2463 – 2947 | 1534 – 2951 |  | 1355 – 2138 | 939 – 1898 |  |
| *Wilcoxon rank sum test. Treatment response was defined as a ≥ 30% reduction in average pain intensity at the 2-month follow-up.*  **p < 0.05* | | | | | | |

| **Table S7.** Relationship between 100-unit volume increases and pain intensity in the methadone group | | | | | |
| --- | --- | --- | --- | --- | --- |
|  | Left | | | Right | |
| Region | N | Beta (IC95%) | p-value | Beta (IC95%) | p-value |
| Insula | 15 | 0.9 (-1.5, 3.3) | 0.454 | 0.9 (-1.4, 3.1) | 0.428 |
| Precentral | 15 | 0.6 (-0.1, 1.3) | 0.095 | 0.4 (-0.2, 1.1) | 0.178 |
| Post-central | 15 | 0.1 (-0.9, 1.2) | 0.820 | -0.1 (-1.1, 0.9) | 0.792 |
| Paracentral | 15 | 1.0 (-1.5, 3.4) | 0.397 | 1.3 (-0.7, 3.3) | 0.192 |
| Caudal anterior cingulum | 15 | -1.0 (-3.5, 1.6) | 0.424 | 2.9 (-2.1, 7.9) | 0.228 |
| Isthmus | 15 | 2.3 (-1.4, 6.1) | 0.205 | 3.9 (0.0, 7.8) | 0.050 |
| Posterior Cingulum | 15 | 0.5 (-2.8, 3.9) | 0.727 | 3.0 (-0.3, 6.3) | 0.069 |
| Rostral anterior cingulum | 15 | 1.4 (-1.3, 4.0) | 0.274 | 2.8 (-1.3, 7.0) | 0.165 |
| *Relationship between increasing cortical volume and estimated VAS at day 56, adjusted for VAS at baseline.*  **p < 0.05* | | | | | |

| **Table S8.** Cortical volume of different brain regions in the placebo group according to treatment response | | | | | | |
| --- | --- | --- | --- | --- | --- | --- |
|  | L | | | R | | |
| Region | Non-responder N = 14 | Responder  N = 8 | p-value | Non-responder N = 14 | Responder  N = 8 | p-value |
| Insula |  |  | 0.4 |  |  | >0.9 |
| Median (Q1, Q3) | 6,626 (5,815, 6,906) | 6,416 (5,906, 6,567) |  | 6,226 (5,608, 6,729) | 6,327 (5,529, 6,624) |  |
| Min – Max | 5,162 – 7,222 | 5,098 – 7,204 |  | 2,509 – 7,110 | 5,227 – 7,271 |  |
| Precentral |  |  | 0.070 |  |  | 0.11 |
| Median (Q1, Q3) | 11,166 (10,584, 13,072) | 9,853 (8,581, 11,240) |  | 10,866 (9,565, 11,902) | 9,515 (9,081, 10,699) |  |
| Min – Max | 9,335 – 14,252 | 8,081 – 13,242 |  | 9,155 – 13,462 | 8,936 – 14,182 |  |
| Postcentral |  |  | 0.11 |  |  | 0.5 |
| Median (Q1, Q3) | 8,290 (7,571, 8,827) | 7,621 (7,007, 7,953) |  | 8,023 (7,337, 8,586) | 7,709 (6,870, 8,309) |  |
| Min – Max | 6,654 – 11,883 | 6,470 – 9,779 |  | 5,697 – 11,101 | 5,750 – 9,702 |  |
| Paracentral |  |  | 0.2 |  |  | 0.003* |
| Median (Q1, Q3) | 3,033 (2,713, 3,388) | 2,675 (2,410, 2,910) |  | 3,450 (3,034, 3,688) | 2,791 (2,709, 2,945) |  |
| Min - Max | 2,343 - 3,782 | 2,239 - 3,987 |  | 2,849 - 3,952 | 2,635 - 3,534 |  |
| Caudal anterior cingulate |  |  | 0.11 |  |  | 0.8 |
| Median (Q1, Q3) | 1,551 (1,332, 1,843) | 1,288 (952, 1,484) |  | 1,570 (1,432, 2,135) | 1,623 (1,325, 1,961) |  |
| Min - Max | 747 - 2,326 | 844 - 1,607 |  | 1,234 - 3,049 | 857 - 2,218 |  |
| Cingulate isthmus |  |  | 0.2 |  |  | 0.3 |
| Median (Q1, Q3) | 2,384 (2,313, 2,944) | 2,162 (2,052, 2,415) |  | 2,154 (2,040, 2,452) | 1,964 (1,921, 2,166) |  |
| Min - Max | 1,651 - 3,472 | 1,748 - 2,526 |  | 1,616 - 2,568 | 1,895 - 2,624 |  |
| Posterior cingulate |  |  | 0.4 |  |  | 0.4 |
| Median (Q1, Q3) | 2,701 (2,553, 2,984) | 2,410 (2,283, 2,882) |  | 2,734 (2,364, 3,171) | 2,490 (2,364, 2,873) |  |
| Min - Max | 1,623 - 3,193 | 1,982 - 3,365 |  | 1,275 - 3,360 | 2,222 - 3,074 |  |
| Rostral anterior cingulate |  |  | 0.2 |  |  | 0.7 |
| Median (Q1, Q3) | 2,229 (1,903, 2,814) | 2,037 (1,668, 2,401) |  | 1,586 (1,275, 2,002) | 1,661 (1,306, 1,806) |  |
| Min - Max | 1,766 - 3,048 | 849 - 3,004 |  | 1,074 - 2,499 | 1,098 - 1,937 |  |
| *Treatment response was defined as a ≥ 30% reduction in average pain intensity at the 2-month follow-up.*  **p < 0.05* | | | | | | |

| **Table S9.** Cortical volume comparison between hemispheres according to brain regions and pain location | | | |
| --- | --- | --- | --- |
| Region | Contralateral  N = 21 | Ipsilateral  N = 21 | p-value |
| Insula | 6,403 (5,665, 6,584) | 6,560 (5,877, 6,762) | 0.781 |
| Precentral | 10,670 (9,271, 11,902) | 11,061 (9,963, 12,521) | 0.035 |
| Post-central | 8,012 (7,082, 8,560) | 7,708 (7,337, 8,692) | 0.862 |
| Paracentral | 3,310 (2,789, 3,648) | 3,137 (2,809, 3,740) | 0.424 |
| Caudal anterior cingulate | 1,559 (1,294, 1,756) | 1,686 (1,406, 1,863) | 0.862 |
| Cingular isthmus | 2,417 (2,040, 2,567) | 2,370 (2,147, 2,545) | 0.741 |
| Posterior cingulate | 2,684 (2,298, 3,071) | 2,771 (2,482, 2,908) | 0.958 |
| Rostral anterior cingulate | 2,071 (1,565, 2,463) | 1,766 (1,355, 2,042) | 0.224 |
| *Cortical volume (Median, Q1, Q3) from ipsilateral and contralateral brain regions, according to laterality of the pain syndrome. Volunteers with bilateral pain were excluded from this analysis (N=21).*  **p < 0.05* | | | |
